# USE OF MACHINE LEARNING TO PREDICT INDIVIDUAL POSTPRANDIAL GLYCEMIC RESPONSES TO FOOD AMONG INDIVIDUALS WITH TYPE 2 DIABETES IN INDIA

**DOI:** 10.1101/2024.02.08.24302542

**Authors:** Niteesh K. Choudhry, Shweta Priyadarshini, Jaganath Swamy, Mridul Mehta

**Author notes:** **Corresponding Author:** Niteesh K. Choudhry, MD, PhD.

## Abstract

**Background:** Type 2 diabetes (DM2) is a leading cause of premature morbidity and mortality globally and affects more than 100 million people in the world’s most populous country, India. Nutrition is a critical and evidence-based component of effective blood sugar control and most dietary advice emphasizes carbohydrate and calorie reduction. Emerging global evidence demonstrates marked inter-individual differences in post-prandial glucose response (PPGR) although no such data exists in India and prior studies have primarily evaluated PPGR variation in individuals without diabetes.

**Methods:** This prospective cohort study seeks to characterize the PPGR variability in Indians with diabetes and to identify factors associated with these differences. Adults with type 2 diabetes and a hemoglobin A1c ≥7 are being enrolled from 14 sites around India. Subjects wear a continuous glucose monitor, eat a series of standardized meals, and record all free-leaving foods, activities, and medication use for a 14-day period. The study’s primary outcome is PPGR, calculated as the incremental area under the curve 2 hours after each logged meal.

**Discussion:** This study will provide the first large scale examination variability in blood sugar responses to food in India and will be among the first to estimate PPGR variability for individuals with DM2 in any jurisdiction. Results from our study will generate data to facilitate the creation of machine learning models to predict individual PPGR responses and to facilitate the prescription of personalized diets for individuals with DM2.

---

Type 2 diabetes mellitus (DM2) is the leading cause of chronic kidney disease, end-stage renal disease, blindness and non-traumatic amputation; it also substantially increases the risk of myocardial infarction, stroke and heart failure.^1^ Its prevalence is particularly high in India, which is now the most populous country in the world. As of 2023, more than 100 million Indians have diabetes, representing more than 11% of the population.^2^ An additional 136 million Indians have prediabetes. These numbers are anticipated to continue to grow rapidly. The lifetime risk of DM2 among obese 20-year-old Indians is estimated to be more than 86%.^3^ The rising prevalence of this condition in India is believed to be the result of changing diets, increasingly sedentary occupations, lower levels of physical activity in the context of urbanization, and rapidly increasing rates of obesity.^4^

These trends are particularly concerning because of important differences between the presentation and consequences in DM2 in Indians compared with other racial and ethnic groups. These differences, which often referred to as the “Asian Indian Phenotype”,^5–7^ are characterized by the onset of DM2 at a younger age and substantially lower body-mass index (BMI) than people of other races and ethnicities.^8–10^ Indians have higher levels of insulin resistance (and for longer periods of time) and premature beta-cell failure.^7^ They are more likely to develop the fatal complications of DM2, most notably heart disease.^7^ These features are thought to result from a mix of lifestyle, epigenetics, and fetal programming factors.^7, 11, 12^

The fundamental goal of diabetes management is to maintain near-normal glucose levels. A variety of self-management behaviors, in particular adherence to diet and regular exercise, are central to this goal. An extensive body of evidence demonstrates that aiding DM2 patients with self-management behaviors is associated with improvements in a wide range of outcomes including knowledge, self-care behaviors, weight, quality of life, hemoglobin A1c (A1c), all-cause mortality and health care costs.^13, 14^

Guidelines recommend that nutritional guidance be personalized based on nutritional status, lifestyle, and metabolic goals.^15^ Despite this, most dietary advice for individuals with DM2 remains generic emphasizing reductions in calories and minimization of carbohydrates.^16^ However, there are marked inter-individual responses in post-prandial glucose response (PPGR).^17^ A study conducted in Israel found substantial PPGR variability to standardized meals for individuals without diabetes.^18^ Similar data has been generated in the UK, US and China.^19–21^

There have been no studies characterizing food responsiveness among Indians and virtually no published data, from any judication, in the variability in PPGR for individuals with DM2.^22^ Given the unique Indian diabetes phenotype and differences between Indian and western diets, there are very likely to be differences in blood sugar responses to food and exercise in India than observed elsewhere, just as there have been in Indians’ responses to diabetes medications.^25^ Accordingly, the goal of this study was to characterize and identify factors associated the variability in PPGR among individuals with DM2 in India.

## METHODS

This prospective cohort study seeks to evaluate the relationship between PPGR and self-management activities including diet, exercise, and other daily routines, for individuals with DM2 in India. This study was approved by the ethics committees at all institutions enrolling patients and is registered with Clinical Trials Registry-India (CTRI/2022/02/040619). The authors are responsible for the design, conduct and analysis of this study and all met International Committee of Medical Journal Editors (ICJME) criteria.

### Study setting

This trial is being conducted at 14 outpatient clinics in geographically distinct regions across India. Sites were identified and managed by a multinational contract research organization and were included if they specialized in the care of individuals with diabetes, had an established research infrastructure for the conduct of diabetes-related studies including a Site Principal Investigator who is a diabetologist (with clinical training in Endocrinology or Internal Medicine) and a local ethics committee to provide study oversight, and a sufficient volume of potentially-eligible patients. Study enrollment began in May 2022.

### Eligible subjects and enrollment

The study population consists of adults with diabetes and suboptimal disease control, classified A1c ≥7%. Complete inclusion and exclusion criteria are summarized in **Table 1**.

**TABLE 1:** Subject inclusion and exclusion criteria.

|  |  |
| --- | --- |
| Inclusion<br>Criteria | ▪ Age $\geq 18$ and $<75$ years<br>▪ Physician-diagnosed type 2 diabetes treated with $\geq 1$ oral hypoglycemic agents (concurrent treatment with an injectable non-insulin agent is permitted)<br>▪ Hemoglobin A1c (HbA1c) $\geq 7.0\%$ recorded within the past 30 days<br>▪ Mobile phone capable of running the protocol-specified applications<br>▪ Functional English literacy |
| Exclusion<br>Criteria | ▪ Unable or unwilling to provide informed consent or comply with the study-specified procedures<br>▪ Current use of prandial insulin including a continuous insulin infusion pump<br>▪ Currently pregnant or planning to become pregnant<br>▪ Estimated life expectancy $\leq 12$ months<br>▪ Active cancer<br>▪ Myocardial infarction or stroke in the last 6 months<br>▪ Receiving or planned to initiate dialysis for end-stage renal disease<br>▪ Receiving oral or intravenous steroids<br>▪ Any contraindication to using a continuous glucose monitor (CGM) |

Potentially eligible patients are identified from clinic records and are invited to attend an in-person screening visit at which time eligibility is confirmed and written informed consent obtained. Consenting patients are asked to provide sociodemographic and medical information (specifically, age, sex, predominant diet, health conditions, family history, and current medications) and to complete baseline surveys including the World Health Organisation STEPwise Approach to NCD Risk Factor Surveillance (STEPS) survey,^23^ World Health Organisation-Five Well-Being Index (WHO-5),^24^ Diabetes Distress Scale,^25^ Wilson Adherence Scale^26^ and the Pittsburgh Sleep Quality Index.^27^

Biometric data including blood pressure, heart rate, weight, height, and body measurements at the upper arm, thigh, calf, waist, and hips, are collected by study coordinators at each site. Finally, enrolled subjects provide blood samples including a complete blood count, hemoglobin A1c, blood electrolytes, creatinine, and cholesterol, as well as a urinalysis.

After completing baseline assessments, subjects are fitted with an Abbott Freestyle Libre continuous glucose monitors (CGM) sensor on their upper, non-dominant arm and are provided with a Xiaomi Mi Band Smart Wristband (heart rate monitor) and a Roche Accu-Chek glucometer with testing supplies, and dietary supplements to be consumed with their standardized meals (see **Follow-up procedures** below). Study-specific applications are downloaded on to the subjects’ smartphones to allow them to log dietary intake and synchronize their continuous glucose and heart rate monitors. As a back-up, subjects are given a paper dietary logbook and a Freestyle Libre CGM reader with which to collect protocol-specified data.

### Follow-up procedures

Subjects are followed for 14 days. They are instructed to wear the CGM and heart rate monitor. The heart rate monitor is to be always worn, including during sleep, and only removed for re-charging. Subjects are also asked to check their capillary glucose on days 2 through 6 before breakfast and dinner.

Subjects log their full dietary intake using the study app or logbook over the 14-day study period, including all standardized test meals and free-living foods (including snacks), beverages (including water) and medications. Participants also log all exercise.

Subjects are required to consume protocol-specified meals and to perform light activity, as described in **Table 2**. The standardized meals consist of vegetarian breakfast foods which subjects are to prepare in their homes. The meals vary in their proportion of carbohydrate, fiber, protein, and fat (see **Table 3**). Subjects are instructed to: (a) fast for a minimum of 8 hours prior to and 3 hours after consuming the standardized breakfast meal; (b) during these fasting periods, limit exercise and drink only still (not sparkling) water, tea or coffee in moderation; (c) eat the meal, in its entirety, within 20 minutes. After completing the post-meal fasting period on standardized test meal days, subjects may consume other foods as they normally would unless there are other meal modifications specified by the protocol later the same day.

**TABLE 2:** Meal schedule during the at-home study period.

| **Day** | **Timing** | **Meal type** |
| --- | --- | --- |
| **1** | Breakfast | Fasting |
|  | Lunch | As desired |
|  | Dinner | As desired |
| **2** | Breakfast | Perceived healthy meal |
|  | Lunch | Post meal exercise |
|  | Dinner | Typical mixed protein with dinner |
| **3** | Breakfast | Fiber supplement with standardized test meal |
|  | Lunch | Pre-meal exercise |
|  | Dinner | Alternative mixed protein with dinner |
| **4** | Breakfast | Fiber supplement prior to standardized test meal |
|  | Lunch | As desired |
|  | Dinner | As desired |
| **5** | Breakfast | Protein supplement with standardized test meal |
|  | Lunch | As desired |
|  | Dinner | Alternative mixed protein with dinner |
| **6** | Breakfast | Protein supplement prior to standardized test meal |
|  | Lunch | Added fiber with lunch |
|  | Dinner | Alternative mixed protein with added protein |
| **7** | Breakfast | Regular breakfast |
|  | Lunch | Added fruit |
|  | Dinner | Cooled carbohydrate |
| **8** | Breakfast | Regular breakfast with added protein |
|  | Lunch | Added fruit with protein |
|  | Dinner | Regular carbohydrate |
| **9** | Breakfast | Protein followed by carbohydrate |
|  | Lunch | Water 30 minutes before lunch |
|  | Dinner | As desired |
| **10** | Breakfast | Standardized test meal |
|  | Lunch | As desired including dessert |
|  | Dinner | Protein followed by carbohydrates and vegetables |
| **11** | Breakfast | Standardized test meal with post-meal exercise |
|  | Lunch | As desired including desert and cinnamon |
|  | Dinner | Vegetables followed by protein then carbohydrates |
| **12** | Breakfast | Standardized test meal with post-meal exercise |
|  | Lunch | As desired |
|  | Dinner | As desired finishing eating by 8pm |
| **13** | Breakfast | Water before late breakfast + post-meal exercise |
|  | Lunch | Low glycemic index lunch |
|  | Dinner | Low glycemic index dinner + finish dinner by 8p + exercise after meal |
| **14** | Breakfast | Water before late breakfast + exercise after meal |
|  | Lunch | Low glycemic index lunch + exercise after meal |
|  | Dinner | Low glycemic index dinner + finish dinner by 8p + exercise after meal |

**TABLE 3:** Nutritional composition of standardized test meals.

| **Meal type** | **Example meal** | **Energy**<br>**(kcal)** | **Carbohydrate**<br>**(g)** | **Fat**<br>**(g)** | **Protein**<br>**(g)** | **Fiber**<br>**(g)** | **Carbohydrate**<br>**(%energy)** | **Fat**<br>**(%energy)** | **Protein**<br>**(%energy)** |
| --- | --- | --- | --- | --- | --- | --- | --- | --- | --- |
| Carbohydrate | 2 aloo paratha with curd | 474.4 | 61.5 | 12.5 | 7.7 | 8.3 | 51.9 | 23.8 | 6.46 |
| Added fiber | 2 aloo paratha with curd<br>+ fiber supplement | 506.4 | 64.1 | 12.5 | 7.8 | 21.9 | 50.6 | 22.3 | 6.05 |
| Added protein | 2 aloo paratha with<br>curd + protein<br>supplement | 594.4 | 62.9 | 14.6 | 31.9 | 8.3 | 42.3 | 22.1 | 21.4 |

On other days, subjects are asked to consume normal foods with protocol-specified constraints. For example, on different days, subjects vary the types of mixed protein (e.g., different types of lentils with or without added protein), ordering with which foods are consumed (e.g., protein before carbohydrate v. protein with carbohydrate), consume water before their meal, go for a walk after eating or eat what they perceive to be a healthy meal. Where applicable, subjects are given several options as to which of their usual foods are acceptable for each protocol specified food modification.

During the follow-up period subjects are contacted by phone and text messages to ensure protocol compliance. If the outreach identifies a problem with a CGM (i.e., it was damaged, fell off or malfunctioned), study staff provide a replacement within 24 hours during which time subjects are asked to pause their meal protocol and restart once their CGM has been re-applied and recalibrated. If a test meal was not consumed as intended, participants are provided with the option to repeat the meal.

On Day 15, subjects are asked to remove their CGM and will complete endline surveys. If need be, study staff help with recording into the study app or logbook of missing or inaccurate food, activity, and medication data.

### Statistical analysis plan

The study’s primary outcome is PPGR. Following the Wolever and Jenkins method,^28^ as adapted by Zeevi et al,^18^ Mendes-Soares et al^20^ and Berry et al,^29^ logged meal times and continuous glucose measurements will be used to calculate the incremental area under the curve (iAUC). Prior to conducting analyses, meals logged less than 30 minutes apart will be merged and meals logged within 90 min of other meals will be removed. Very small (<15 g and <70 calories) meals and meals with very large (>1 kg) components, meals with incomplete logging and meals consumed at the first and last 12 hour of the CGM connection will also be removed. To reduce noise, the median of all glucose values from the 30-minute period prior to the meal will be taken as the initial glucose level, above which the incremental area will be calculated. Meals that had incomplete glucose measurements in the time window of 30 minutes before and 2 hours after the logged mealtime will be filtered out.

Descriptive statistics will be used to plot the range of PPGRs responses to standardized test meals as well as the correlation between PPGR and the nutritional composition of the logged meals (i.e., carbohydrates, fat, and protein).

A machine learning predictor will be developed based on stochastic gradient boosting regression (XGBoost, version 0.6)^30^ using the XGBRegressor class. Postprandial glycemic responses will be predicted as the sum of predictions from thousands of decision trees. Trees will be inferred sequentially, with each trained on the residual of all previous trees and making a small contribution to the overall prediction. The features incorporated in each tree are selected by an inference procedure from a pool of features representing meal content, demographics, health habits, baseline laboratory values, as well as CGM, heart rate and activity data.

Performance will be assessed by 5-fold cross-validation in which participants are divided into 10 groups, the model will be trained on 9 parts, and the performance will be measured by the ability to accurately predict meals reported by the left-out participants (out-of-bag predictions). Prediction results on subjects from all left folds will be aggregated, and Pearson product moment correlation with the measured post-prandial glucose responses will be reported. The standard error for the calculated performance will be assessed using 50 iterations of bootstrapping. Random data sets of the same size as the original will be sampled with replacement from the original data set, and the entire training and validation process was repeated.

### Sample size considerations

1050 individuals will be targeted for recruitment. Assuming a 5% loss-to-follow-up, this corresponds to 1000 evaluable individuals at the end of the study. The study has been designed to predict postprandial glucose responses based on individual characteristics and 1000 subjects followed for 14-days will result more than 4 million glucose readings (assuming glucose readings from the CGMs every 5 minutes) and 42,000 meals (assuming 3 meals per participant per day). This volume of data will also provide more than 80% power to detect correlations of a magnitude of r=0.13 (R^2^=0.017) with an p<0.005. We will also be sufficiently powered to detect effects of r=0.165 (R^2^=0.027, i.e., explaining 2.7% of interindividual variation) with p<0.00001, i.e., accounting for 5000 independent hypothesis tests.

## DISCUSSION

The prevalence of DM2 has grown increased rapidly in India such that 1 in 6 people in the word with this condition live in this country.^31^ Along with medications and physical activity, diet is a key tenant of effective blood sugar control.^15^ Guidelines call for individualization of meal planning, which is sometimes referred to as “Medical Nutritional Therapy”. Despite this, personalization of dietary plans are generally based upon broad constructs like age, activity level, health status and preferences, and, for all patients, tend to emphasize overall calorie reductions and minimization of carbohydrate,^16^ especially added sugars and refined grains, in favor of the consumption of non-starchy vegetables and foods that are high in protein.^32^

However, emerging data demonstrates that there are marked inter-individual responses to food,^17^ attributed to differences in physical activity,^33^ gut microbiome,^18, 19, 34^ and genetics^35^ including in variations in skeletal glucose transporters related to insulin resistance.^36^ For example, a study conducted in the US among non-diabetic individuals with a mean BMI of 27 found PPGR to a standardized meal of bagel and cream cheese ranged from 6 to 94 mg/dL.^20^ A similar study conducted in Israel enrolled non-diabetic individuals of whom three-quarters had a BMI ≥ 25 and found mean PPGR to bread and butter of 44 mg/dL*hour but the bottom decile had responses of ≤ 15 mg/dL*hour and the top decile has responses ≥ 79 mg/dL*hour.^18^ Similar data has been generated for individual without diabetes in the UK and US^19, 20^ and for individuals with type 1 diabetes in Israel.^37^

There is exceptionally limited data for variability in PPGR for individuals with DM2 in the peer-reviewed although it is highly likely that such variability exists.^22^ The primary goal of our study is to fill this void and to generate an India-specific machine-learning models on the basis of which PPGR can be predicted with high accuracy for DM2. Similar models have been built in other jurisdictions. For example, a machine learning algorithm trained on CGM data, dietary, activity, anthropometrics and gut microbiota for non-diabetic individuals in Israel was much more accurate at predicting PPGR than generic models based on the carbohydrate content or the amount of calories in a meal.^18^ A separate US based study had similar findings.^20^

Among individuals with diabetes, a study in the Netherlands that included a small number of individuals with DM2 along with individuals with pre-diabetes and normal glucose metabolism, a machine learning model based on CGM data was highly accurate and predicting future glucose values but this study did not specifically evaluate the ability to predict PPGR.^38^ A US study of 1,000 patients of whom one-quarter had DM2 found that a machine learning model trained on CGM, HRM data and food logs was highly accurate at predicting PPGR but this study has, to our knowledge, only been published in abstract form.^22^ These studies have all relied on CGM data to make their predictions. While these devices are increasingly used, practice guidelines do not recommend their long-term use for most individuals with DM2.^39^ Accordingly, a key goal of our study will be to explore the ability to predict PPGR response without reliance on CGM data or with very limited blood sugar data from patients.

There are several limitations to our approach. Our approach is purposely pragmatic and is intended to simulate real-world circumstances for individuals with DM2 living in India. Similar to studies conducted in other jurisdictions, we rely on self-reported dietary and activity information. And, while we are auditing patient logs on an ongoing basis, there may nevertheless be issues with protocol adherence that may undermine the accuracy of the data we collect. Subjects are being recruited from clinics predominantly caring for individuals with diabetes, are required to have functional English literacy and a cellphone capable of running study specific devices. Thus, our results may not be fully generalizable to patients who do not fulfill these criteria. Finally, some of enrollment has overlapped with the COVID-19 pandemic which may have influenced access to health care, dietary practices and glucose control for individuals with DM2.

In conclusion, this study will provide the first large scale examination variability in blood sugar responses to food in India and will be among the first to estimate PPGR variability for individuals with DM2 in any jurisdiction. Results from our study will generate data to facilitate the creation of machine learning models to predict individual PPGR responses and to ultimately facilitate the prescription of truly personalized diets for individuals with DM2.

**FIGURE 1:**
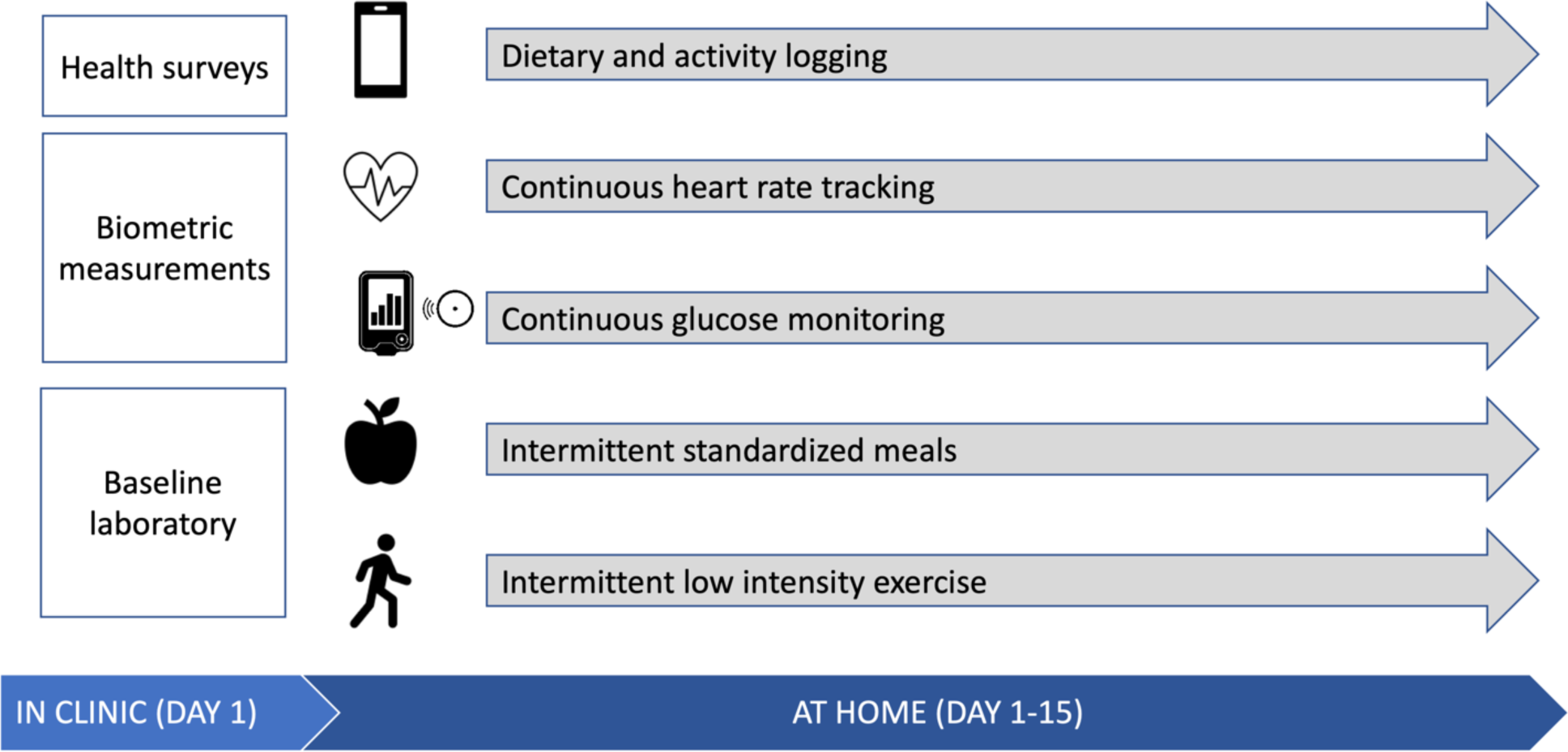
Overall study design.

## Data Availability

All data produced in the present study are available upon reasonable request to the authors

## REFERENCES

1. Pratley RE. The early treatment of type 2 diabetes. Am J Med. Sep 2013;126(9 Suppl 1):S2–9. doi:10.1016/j.amjmed.2013.06.007

2. Anjana RM, Unnikrishnan R, Deepa M, et al. Metabolic non-communicable disease health report of India: the ICMR-INDIAB national cross-sectional study (ICMR-INDIAB-17). Lancet Diabetes Endocrinol. Jul 2023;11(7):474–489. doi:10.1016/S2213-8587(23)00119-5

3. Luhar S, Kondal D, Jones R, et al. Lifetime risk of diabetes in metropolitan cities in India. Diabetologia. Mar 2021;64(3):521–529. doi:10.1007/s00125-020-05330-1

4. India State-Level Disease Burden Initiative Diabetes C. The increasing burden of diabetes and variations among the states of India: the Global Burden of Disease Study 1990-2016. Lancet Glob Health. Dec 2018;6(12):e1352–e1362. doi:10.1016/S2214-109X(18)30387-5

5. Patel SA, Shivashankar R, Ali MK, et al. Is the “South Asian Phenotype” Unique to South Asians?: Comparing Cardiometabolic Risk Factors in the CARRS and NHANES Studies. Glob Heart. Mar 2016;11(1):89–96 e3. doi:10.1016/j.gheart.2015.12.010

6. Shariff AI, Kumar N, Yancy WS, Jr., Corsino L. Type 2 Diabetes and Atherosclerotic Cardiovascular Disease in South Asians: a Unique Population with a Growing Challenge. Curr Diab Rep. Jan 30 2020;20(1):4. doi:10.1007/s11892-020-1291-6

7. Sattar N, Gill JM. Type 2 diabetes in migrant south Asians: mechanisms, mitigation, and management. Lancet Diabetes Endocrinol. Dec 2015;3(12):1004–16. doi:10.1016/S2213-8587(15)00326-5

8. Ramachandran A, Snehalatha C, Kapur A, et al. High prevalence of diabetes and impaired glucose tolerance in India: National Urban Diabetes Survey. Diabetologia. Sep 2001;44(9):1094–101. doi:10.1007/s001250100627

9. Qiao Q, Hu G, Tuomilehto J, et al. Age- and sex-specific prevalence of diabetes and impaired glucose regulation in 11 Asian cohorts. Diabetes Care. Jun 2003;26(6):1770–80. doi:10.2337/diacare.26.6.1770

10. Hsu WC, Araneta MR, Kanaya AM, Chiang JL, Fujimoto W. BMI cut points to identify at-risk Asian Americans for type 2 diabetes screening. Diabetes Care. Jan 2015;38(1):150–8. doi:10.2337/dc14-2391

11. Unnikrishnan R, Gupta PK, Mohan V. Diabetes in South Asians: Phenotype, Clinical Presentation, and Natural History. Curr Diab Rep. Apr 18 2018;18(6):30. doi:10.1007/s11892-018-1002-8

12. Gujral UP, Pradeepa R, Weber MB, Narayan KM, Mohan V. Type 2 diabetes in South Asians: similarities and differences with white Caucasian and other populations. Ann N Y Acad Sci. Apr 2013;1281:51–63. doi:10.1111/j.1749-6632.2012.06838.x

13. Powers MA, Bardsley J, Cypress M, et al. Diabetes Self-management Education and Support in Type 2 Diabetes. Diabetes Educ. Feb 2017;43(1):40–53. doi:10.1177/0145721716689694

14. American Diabetes A. 5. Facilitating Behavior Change and Well-being to Improve Health Outcomes: Standards of Medical Care in Diabetes-2021. Diabetes Care. Jan 2021;44(Suppl 1):S53–S72. doi:10.2337/dc21-S005

15. ElSayed NA, Aleppo G, Aroda VR, et al. 5. Facilitating Positive Health Behaviors and Well-being to Improve Health Outcomes: Standards of Care in Diabetes-2023. Diabetes Care. Jan 1 2023;46(Supple 1):S68–S96. doi:10.2337/dc23-S005

16. Anjana RM, Srinivasan S, Sudha V, et al. Macronutrient Recommendations for Remission and Prevention of Diabetes in Asian Indians Based on a Data-Driven Optimization Model: The ICMR-INDIAB National Study. Diabetes Care. Aug 18 2022;doi:10.2337/dc22-0627

17. Vega-Lopez S, Ausman LM, Griffith JL, Lichtenstein AH. Interindividual variability and intra-individual reproducibility of glycemic index values for commercial white bread. Diabetes Care. Jun 2007;30(6):1412–7. doi:10.2337/dc06-1598

18. Zeevi D, Korem T, Zmora N, et al. Personalized Nutrition by Prediction of Glycemic Responses. Cell. Nov 19 2015;163(5):1079–1094. doi:10.1016/j.cell.2015.11.001

19. Asnicar F, Berry SE, Valdes AM, et al. Microbiome connections with host metabolism and habitual diet from 1,098 deeply phenotyped individuals. Nat Med. Feb 2021;27(2):321–332. doi:10.1038/s41591-020-01183-8

20. Mendes-Soares H, Raveh-Sadka T, Azulay S, et al. Assessment of a Personalized Approach to Predicting Postprandial Glycemic Responses to Food Among Individuals Without Diabetes. JAMA Netw Open. Feb 1 2019;2(2):e188102. doi:10.1001/jamanetworkopen.2018.8102

21. Ma Y, Fu Y, Tian Y, et al. Individual Postprandial Glycemic Responses to Diet in n-of-1 Trials: Westlake N-of-1 Trials for Macronutrient Intake (WE-MACNUTR). J Nutr. Oct 1 2021;151(10):3158–3167. doi:10.1093/jn/nxab227

22. Dalal P, Yazdani M, Snyder M, Rahili S, Torbaghan SS. 75-LB: Machine-Learned, Biophysical Prediction of Glucose Response for 1,000 Subjects. Diabetes. 2020;69(Supplement_1)doi:10.2337/db20-75-LB

23. World Health Organization. STEPwise Approach to NCD Risk Factor Surveillance (STEPS). https://www.who.int/teams/noncommunicable-diseases/surveillance/systems-tools/steps

24. Topp CW, Ostergaard SD, Sondergaard S, Bech P. The WHO-5 Well-Being Index: a systematic review of the literature. Psychother Psychosom. 2015;84(3):167–76. doi:10.1159/000376585

25. Polonsky WH, Fisher L, Earles J, Dudl RJ, Lees J, Mullan J, Jackson RA. Assessing psychosocial distress in diabetes: development of the diabetes distress scale. Diabetes Care. Mar 2005;28(3):626–31. doi:10.2337/diacare.28.3.626

26. Wilson IB, Lee Y, Michaud J, Fowler FJ, Jr., Rogers WH. Validation of a New Three-Item Self-Report Measure for Medication Adherence. AIDS Behav. Nov 2016;20(11):2700–2708. doi:10.1007/s10461-016-1406-x

27. Pittsburgh Sleep Quality Index (PSQI). https://eprovide.mapi-trust.org/instruments/pittsburgh-sleep-quality-index#basic_description

28. Wolever TM, Jenkins DJ. The use of the glycemic index in predicting the blood glucose response to mixed meals. Am J Clin Nutr. Jan 1986;43(1):167–72. doi:10.1093/ajcn/43.1.167

29. Berry SE, Valdes AM, Drew DA, et al. Human postprandial responses to food and potential for precision nutrition. Nat Med. Jun 2020;26(6):964–973. doi:10.1038/s41591-020-0934-0

30. Chen T, Guestrin C. XGBoost: A Scalable Tree Boosting System. presented at: Proceedings of the 22nd ACM SIGKDD International Conference on Knowledge Discovery and Data Mining; 2016; San Francisco, California, USA. 10.1145/2939672.2939785

31. International Diabetes Federation. https://idf.org/our-network/regions-members/south-east-asia/members/94-india.html

32. Evert AB, Dennison M, Gardner CD, et al. Nutrition Therapy for Adults With Diabetes or Prediabetes: A Consensus Report. Diabetes Care. May 2019;42(5):731–754. doi:10.2337/dci19-0014

33. Dunstan DW, Kingwell BA, Larsen R, et al. Breaking up prolonged sitting reduces postprandial glucose and insulin responses. Diabetes Care. May 2012;35(5):976–83. doi:10.2337/dc11-1931

34. Sondertoft NB, Vogt JK, Arumugam M, et al. The intestinal microbiome is a co-determinant of the postprandial plasma glucose response. PLoS One. 2020;15(9):e0238648. doi:10.1371/journal.pone.0238648

35. Carpenter D, Dhar S, Mitchell LM, et al. Obesity, starch digestion and amylase: association between copy number variants at human salivary (AMY1) and pancreatic (AMY2) amylase genes. Hum Mol Genet. Jun 15 2015;24(12):3472–80. doi:10.1093/hmg/ddv098

36. Gibbs EM, Stock JL, McCoid SC, et al. Glycemic improvement in diabetic db/db mice by overexpression of the human insulin-regulatable glucose transporter (GLUT4). J Clin Invest. Apr 1995;95(4):1512–8. doi:10.1172/JCI117823

37. Shilo S, Godneva A, Rachmiel M, et al. Prediction of Personal Glycemic Responses to Food for Individuals With Type 1 Diabetes Through Integration of Clinical and Microbial Data. Diabetes Care. Mar 1 2022;45(3):502–511. doi:10.2337/dc21-1048

38. van Doorn W, Foreman YD, Schaper NC, et al. Machine learning-based glucose prediction with use of continuous glucose and physical activity monitoring data: The Maastricht Study. PLoS One. 2021;16(6):e0253125. doi:10.1371/journal.pone.0253125

39. American Diabetes Association Professional Practice C. 7. Diabetes Technology: Standards of Care in Diabetes-2024. Diabetes Care. Jan 1 2024;47(Suppl 1):S126–S144. doi:10.2337/dc24-S007

